# Health Coaching as a Tool for Lifestyle changes. Insights from the Priomed^®^ Training and Pilot intervention

**DOI:** 10.1101/2025.11.15.25340170

**Authors:** Johanna Hämäläinen, Krista Rapo, Ari Langinkoski, Mikko Rinta, Pekka Aroviita, Ari Haaranen, Julia Kettinen, Mika Venojärvi

**Affiliations:** School of Medicine, Institute of Biomedicine: Sports and Exercise Medicine, University of Eastern Finland; Terveystalo Oyj; Department of Nursing Science, University of Eastern Finland

**Keywords:** lifestyle, coaching, experiences, health promotion

## Abstract

Chronic diseases represent a growing global health burden, accounting for nearly 41 million deaths annually. Beyond their impact on individual health, these conditions place significant strain on healthcare system, increasing societal costs and reducing productivity among working-age population.

Health coaching seems to be a promising method in preventing chronic diseases. However, limited research exists on how health coaching training programs are experienced by health coaches and clients. This study describes the experiences of participants in the PrioMed^®^ health coach training program and a related coaching pilot, aiming to provide insights that can inform future training development and coaching implementation.

This mixed-method study, conducted in collaboration with the Nordic Health Academy, private health care (Terveystalo), and the University of Eastern Finland. The data collection comprised a questionnaire and semi-structured individual interviews. The questionnaire results were summarized using frequency tables, and the interview data were analyzed using inductive content analysis.

The findings indicate that the training program enhanced coaching skills and the insecurity experienced by the coaches was not conveyed to the coaches. Coaches highlighted the importance of practical coaching skills in training, and a healthcare background was considered beneficial for the coaching role. The coachees expressed satisfaction with the coaching methods and would recommend coaching to others.

Coaches provided support, encouragement, and assistance to their coachees.

The study suggests that PrioMed^®^ health coaching effectively addressed individual needs in health behavior change. The findings can inform the development of effective training programs and support the integration of coaching into preventive healthcare.

## INTRODUCTION

Chronic diseases are responsible for approximately 41 million deaths globally each year, accounting for 74% of all deaths. Of these, 17 million are considered premature, occurring before the age of 70 (WHO, 2022), and are significantly affecting the working-age population. More than 80% of premature deaths are caused by cardiovascular diseases, cancers, chronic respiratory diseases and type 2 diabetes (WHO, 2023).

Chronic diseases develop due to a combination of genetic, physiological, environmental, and behavioral factors. Most significant behavioral risk factors include smoking, physical inactivity, unhealthy diet, and harmful alcohol use. (Wharton et al., 2020; WHO, 2023) Metabolic risk factors-such as high blood pressure, overweight and obesity, hyperglycemia, and hyperlipidemia-further increase the risk of mortality and place a growing burden on healthcare system (Chew et al., 2023). However, the development and progression of chronic diseases can be prevented by reducing these risk factors (Wharton et al., 2020; WHO, 2023). Lifestyle choices play a crucial role in both the onset and prevention of chronic diseases (Finnish Institute for Health and Welfare, 2023; Piercy et al., 2018). Since the working-age population is considered to be a productive segment of the national economy, its health impacts not only individuals but also society as a whole (WHO 2023).

One promising approach to addressing these challenges is health coaching, which supports individuals in making sustainable lifestyle changes. Health coaching has been recognized as an effective method for preventing lifestyle-related chronic diseases and managing existing conditions (Carter et al., 2015; Kivelä et al., 2014). Health coaching is defined as a health-focused, goal-oriented process in which the client actively sets personal health goals and, with the support of a coach, works toward achieving them (Olsen, 2014; Wolever et al., 2013). Health coaching is considered a potentially cost-effective strategy for both prevention and disease management. Research indicates that it can positively influence health behaviors and, in doing so, help reduce the rising healthcare costs associated with chronic diseases.

While health coaching has shown promising results in the prevention and management of chronic diseases, the implementation of coaching requires appropriate training.

Health coaching trainees often come from healthcare backgrounds like nursing, particularly when involved in chronic disease treatment programs (Gastala et al., 2018). A nursing background can be advantageous, as communication, a core component of coaching, is already integral to the nursing profession (Obro et al., 2023).

Health coaching training programs are typically short, ranging from a few hours (Davies et al., 2020; Pounds et al., 2015) to a couple of days (Engelhard et al., 2018; Maini et al., 2020). The content of the training is mainly based on the skills needed in coaching, such as motivational interviewing (Pounds et al. 2015, Engelhard et al. 2018, Davies et al.2020), goal setting (Engelhard et al. 2018, Davies et al. 2020, Maini et al. 2020),solution-orientation (Davies et al. 2020, Maini et al. 2020), active listening and reflection (Maini et al. 2020)

Health coaching training has been found to improve participants’ confidence and competence. Up to 90% of participants report understanding coaching techniques, trusting their effectiveness (Davies et al., 2020), and feeling confident in their ability to motivate and support clients in achieving their goals (Pounds et al., 2015). Coaching has been associated with increased professional enjoyment and self-efficacy in the coach role (Engelhard et al., 2018). Despite these positive outcomes, some healthcare professionals initially find adopting the coach role stressful and challenging (Noll et al., 2022). Difficulties in shifting from a traditional expert-based healthcare role to a more client-centered coaching role have been reported, particularly at the beginning of the coaching process (Davies et al., 2020; Maini et al., 2020; Nessen et al., 2014).

Previous studies indicate that clients participating in health coaching are generally satisfied with the process. Clients have reported improvements in both physical and psychological well-being, as well as improved social life. The coach’s role has been found to be significant with reciprocal communication increasing motivation and commitment to achieve set goals (McGill et al., 2018; Miles et al., 2023; Mitchell et al., 2021; Napoles et al., 2019).

PrioMed^®^ health coaching method, developed in Finland and based on the latest research, aims to support individuals in making lifestyle changes (Langinkoski et al., 2024). The aim of this study is to describe the experiences of the health coach students in PrioMed^®^ health coach training and of the coaching as well as the experiences of coachees who have undergone the coaching. A further aim of this study is to produce information that can be utilized in the planning and implementation of health coach training.

## METHODS

### Study Design

This mixed methods study comprises both quantitative and qualitative components. It explores the PrioMed© health coach training program from the perspective of the coaches and the experience of the coaching pilot from the perspective of the coachees. The quantitative component consists of structured questionnaires for the coachees, while the qualitative component includes thematic interviews with both the coaches and the coachees. The qualitative data aimed to provide a deeper understanding of the PrioMed^®^ health coach training program and the coaching pilot function.

The PrioMed^®^ health coach training program was developed using behavior change science, national and EU-level healthcare education guidelines, and structured around the Behavior Change Technique (BCT) taxonomy. The PrioMed^®^ training educated health coaches on behavior change techniques, motivational communication, and needs analysis through online lectures and practical tools. The coaching process was implemented over six months and included seven phone-based sessions per participant. Sessions were highly structured, ranging from 15 to 45 minutes, and personalized using the PrioMed^®^ Lifestyle Questionnaire and Needs Analysis to identify goals (Langinkoski et al. 2024)

### Reseach Setting

This study was conducted in collaboration with Nordic Health Academy, Terveystalo Oyj and the University of eastern Finland. PrioMed^®^ health coaching is developed by Nordic Health Academy, and the coaches were employed at Terveystalo. All study participants were clients of Terveystalo, some referred through the occupational health service, and others self-paying.

### Sample Selection and Recruitment

Experiences from PrioMed^®^ health coaching training program and from the coaching pilot were collected from all coaches (n=4). Experiences from the coaching pilot were collected from those coachees who finished the coaching pilot (n=2). The coachees of the study were recruited from private healthcare (Terveystalo Oyj) customers, and the inclusion criteria included prior participation in an annual health checkup or a digital wellness program. A total of 25 participants completed the study.

### Data Collection

The data used in this study was collected through individual thematic interviews with coaches after completing PrioMed^®^ health coach training and after PrioMed^®^ health coach pilot. Coaches perceived capabilities to work as a health coach were assessed by using a five-staged Likert scale. The data from the coachees consisted of responses to electronic, structured questionnaires sent to coachees who completed the PrioMed^®^ health coach pilot (n=19) and thematic interviews conducted with volunteers (n=5).

Sociodemographic data from the coachees who responded to the questionnaire are presented in Table 1.

**Table 1.** Sociodemographic Characteristics of Survey Respondents.

|  | n | % |
| --- | --- | --- |
| Gender |  |  |
| Woman | 15 | 79 % |
| Man | 4 | 21 % |
| Age (years) |  |  |
| 40–49 | 2 | 11 % |
| 50–59 | 7 | 36 % |
| 60–69 | 2 | 11 % |
| over 70 | 8 | 42 % |
| BMI (kg/m<sup>2</sup>) |  |  |
| Normal, < 25 | 4 | 20 % |
| Overweight, 25–30 | 11 | 58 % |
| Obesity class I, 30–35 | 2 | 11 % |
| Obesity class II, 35–40 | 0 | 0 % |
| Obesity class III, > 40 | 2 | 11 % |
| Marital status |  |  |
| Single-person household | 7 | 37 % |
| Married, no children | 2 | 10 % |
| Married with children | 3 | 16 % |
| Couple, children not living at home | 7 | 37 % |
| Employment status |  |  |
| Full-time employment (more than 35 hours/week) | 10 | 53 % |
| Retired | 9 | 47 % |

### Analysis

The interviews were conducted via Microsoft Teams, and the qualitative data were analyzed by inductive content analysis. The process included open coding, categorization, and abstraction to identify the key themes. Quantitative data from the questionnaire were analyzed using Microsoft Excel.

### Ethics Approval

The study received ethical approval from the ethics committee of the Northern Savo Hospital District, Kuopio. The research was conducted in accordance with the principles of good scientific practice (National board on Research Integrity Tenk, 2023). All participants signed a dated written informed consent form before participating in any of the study assessments.

## RESULTS

### Experiences of the coaches

All the interviewed participants in health coach training were healthcare professionals who had studied either in a university of applied sciences or in secondary education and were now studying in a university of applied sciences. None of the interviewees had received any prior training, such as PrioMed^®^ health coach training before. All had experience in healthcare but no experience in coaching. PrioMed^®^ health coach training was considered good but perceived as too theory-heavy and too short for its scope. The valuable part of the training was the practical exercises, and participants expressed a desire for more of these in the future. Coaches’experiences of PrioMed^®^ health coach training are presented in Table 2.

**Table 2.** Experiences of the PrioMed^®^ health coaching training program.

| What's positive? | To be developed |
| --- | --- |
| • Good lectures and clear diagrams<br>• The information obtained from the training was considered useful<br>• Increasing one's own skills and a new way of thinking<br>• Psychological side of coaching and the realization that changing behavior can be simple when the coachee realizes it themselves<br>• Examples of practical coaching and the exercises that the students were able to participate in themselves<br>• The digital learning environment supported the revision of what had been learned | • Not enough training hours to internalize the amount of information<br>• Lessons were heavy due to the emphasis on theory and the large amount of information<br>• Too many large theoretical entities on behavioural change<br>• Needs more instructions and examples for practical coaching, practical exercises, interactivity and reflection exercises<br>• Needs more condensed materials for the digital learning environment<br>• Coach's manual included in the training-phase |

Although the health coach students felt that the training equipped them with the necessary skills to work as health coaches, all interviewees also expressed some uncertainty about their coaching abilities. A three-week gap between training and the start of coaching increased uncertainty, as participants needed to integrate the newly learned skills. However, prior experience with healthcare consultations and early exposure to coaching calls helped enhance their readiness. After the training, the self-assessed capabilities to work as a health coach in Likert scale were 2-3, which means “little capabilities” or “I don’t know”.

The coaches and the coachees met 5-7 times during the six months. The number of meetings, total length of coaching, and the coaching method were generally considered appropriate, although in some cases prearranged 15-minute length for the session were perceived to be too short. The coaches regarded PrioMed^®^ Lifestyle Questionnaire and PrioMed^®^ Needs Analysis as useful tools. The number of coaches per coach ranged from 7 to 30. As the number of the coaches decreased from four to two during the coaching process, the number of the coachees increased. This change was perceived in different ways; however, the coaches emphasized that the focus should be on scaling the initial phase of coaching rather than on the total number of clients.

The coachees responded well to coaching. Mostly, the change was achieved in 2-4 sessions. In the highly motivated coachees, changes often occurred early in the coaching process. The coaches made efforts to apply the approaches and techniques acquired during the training in their actual coaching. Their use of these was supported by the coach’s manual, sticky notes about phrasal tests and techniques, and peer support from the other coaches. However, the coaches sometimes struggled to recall the full range of techniques and how to apply them, which led to frequent familiar methods. Sometimes, the coachees were reluctant to follow certain processes, requiring the coach to adapt and try to find suitable alternatives.

Challenges in coaching arose when the coachee experiences health problems that required changes to goals and plans when they had longer break in coaching due to the coach’s absence. Another challenge was delay in receiving the PrioMed^®^ Lifestyle Questionnaire and Needs Analysis results at the beginning of the coaching. In addition, the coaches’ concurrent work obligations interfered with coaching, and conversely, the coaching occasionally took time away from their other work responsibilities.

Healthcare education and work experience, particularly conducting nursing appointments and customer calls, were found to be helpful in health coaching. A healthcare background was seen as a strong foundation for understanding the coachees health concerns and implementing health coaching more effectively. At the end of the pilot, the coaches were asked about their capabilities to work as a health coach on Likert scale, and the capability was 4, which means “good capability”.

### Experiences of the coachees

Participants in the PrioMed^®^ health coaching program reported receiving strong support from their coaches, with all respondents (n = 19) indicating encouragement toward personal goals and 73.7% receiving help for their specific concerns. The coaching emphasized physical activity, nutrition, and sleep, which were also the areas where participants felt most supported. Individual needs were addressed through personalized topic selection, flexible goal setting, and session durations tailored to situational requirements. The majority found the number and length of sessions appropriate, and overall satisfaction with the program was high, with 89.5% expressing satisfaction and 84.2% likely to recommend it. (Table 3)

**Table 3.** PrioMed^®^ Health Coaching Program-Results of Questionary.

| Category | Details / Results |
| --- | --- |
| Participants | 19 completed the program and responded to the questionnaire |
| Coach Support | 100% received encouragement and support toward personal goals |
| Help with Specific Issue | 73.7% received help for the issue they sought assistance with |
| Most Discussed Topics | Physical activity: 73.7%<br>Nutrition: 57.9%<br>Sleep: 42.1% |
| Areas of Most Support | Physical activity: 57.9%<br>Nutrition: 52.6%<br>Sleep: 36.8% |
| Interview Insights | Follow-up increased motivation<br>Small steps were emphasized<br>Coaches fostered hope and flexibility |
| Coach Characteristics | Approachable, supportive, clear guidance, flexible goal-setting |
| Recommendation Likelihood | Definitely: 42.1%<br>Maybe: 42.1%<br>Probably not: 5.3% (1 respondent) |
| Personalization | Program considered individual needs; topics chosen by participants |
| Session Quantity Satisfaction | 89.5% found the number of sessions appropriate |
| Session Duration Satisfaction | 94.7% found session length suitable; varied by individual needs |
| Topic Depth | 1–2 topics typically chosen; sufficient for in-depth engagement |
| Program Duration Feedback | Longer duration could allow more topics |
| Method Satisfaction | 94.7% satisfied with coaching methods; 1 respondent dissatisfied |
| Overall Satisfaction | Very satisfied: 31.6%<br>Satisfied: 57.9% |

The interviews highlighted the importance of addressing the most important issue for the coachee, as this often led to improvements in other areas of life. Follow-up sessions were considered meaningful, as they allowed the coachees to review their progress with the coach, and if necessary, explore alternative solutions to challenges. Individual needs were well considered during the coaching process. In particular, the ability to adapt goals and actions when necessary was perceived as positive and supportive. The coachees experiences from PrioMed health coaching can be found in Figure 1.

**Figure 1.**
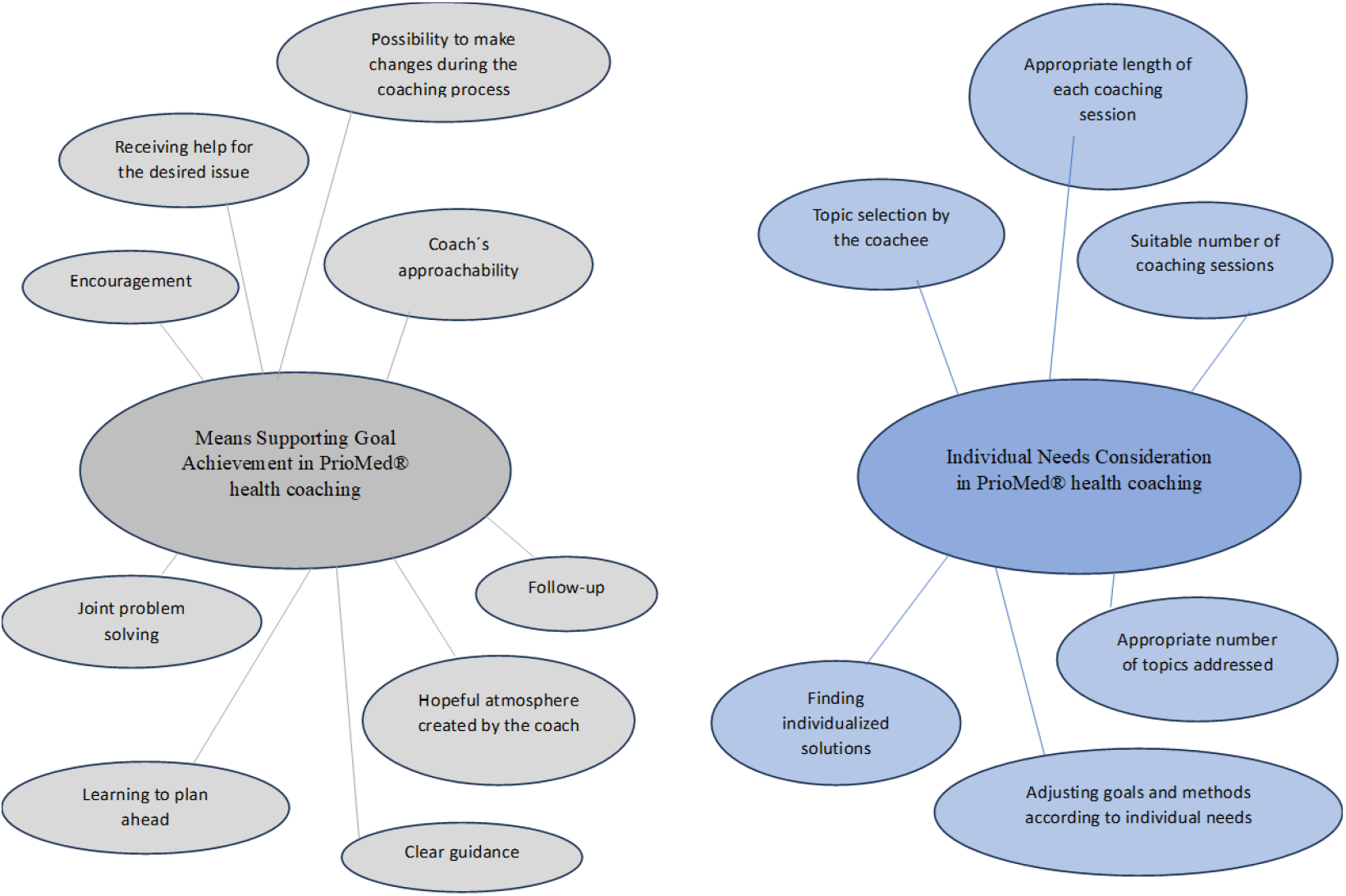
Factors Supporting Goal Achievement and Individual Needs Consideration in PrioMed^®^ Health Coaching

## DISCUSSION

The findings of this study indicate that PrioMed^®^ health coaching training was generally well-received; however, it was considered too brief and overly theoretical relative to the scope of its content. The most appreciated aspect of the program was the practical exercises, which participants wished had been more extensively integrated into the training. Despite post-training uncertainty regarding their readiness, participants reported positive experiences with health coaching. Coachees who took part in the PrioMed^®^ pilot expressed satisfaction with the support they received in making lifestyle changes. They found the coaching methods suitable and responsive to their individual needs. Nearly all participants indicated they would recommend the PrioMed^®^ health coaching program to their friends or close contacts.

Although all participants came from healthcare backgrounds, most had no prior coaching experience and reported that the training’s content and duration did not sufficiently correspond to their initial level of competence. This reflects challenges observed in earlier studies, where the amount of material presented was perceived as excessive (Davies et al., 2020; Noll et al., 2022). While the importance of theoretical knowledge was acknowledged, participants emphasized the need for more opportunities to practice coaching techniques in a realistic context. Practical exercises were considered the most beneficial part of the training and participants expressed a strong desire for more hands-on learning. This aligns with previous research emphasizing the importance of interactive methods and real-world examples for developing coaching competence (Davies et al., 2020; Maini et al., 2020). Coaches also appreciated the psychological aspects of coaching, including the realization that behavior change can be simple when the coachee experiences an “aha” moment, mirroring findings by Maini et al. (2020).

Despite feeling generally prepared, all coaches reported uncertainty regarding their skills and their ability to effectively support clients. This contrasts with previous findings in which participants expressed increased confidence post-training (Davies et al., 2020; Pounds et al., 2015). One contributing factor may have been the time gap between training and actual coaching, which has also been identified as a challenge by Noll et al. (2022). Offering immediate opportunities to apply learned skills could help reinforce confidence and competence.

The coaching process was largely successful. Both coaches and coachees were satisfied with the frequency and content of the sessions, although some coaches felt that 15-minute follow-ups were too brief. From the coachees’ perspective, the sessions were flexible and adapted to their individual needs, indicating that the coaches were able to apply their skills in a client-centered manner. This observation supports previous findings on the importance of tailoring coaching methods to individual preferences (Mitchell et al., 2021; Thom et al., 2015). Coaches reported that the PrioMed^®^ health coaching training was helpful in their coaching practice, and they made efforts to apply the skills they had learned. Support was provided by the training materials, peer discussions among the coaches, and collegial support, which has previously been identified as a key factor for successful implementation (Clason et al., 2023). One recurring challenge was that coaches tended to revert to familiar approaches rather than fully utilizing the new methods introduced in the training. This may indicate a need for more practice-based training and real-life application scenarios to encourage more diverse method use.

Coaching was primarily conducted by phone, and while most participants were satisfied, some expressed a preference for in-person sessions, particularly among those less comfortable with technology. This finding echoes prior research supporting the effectiveness of digital health coaching (Kappes et al., 2023; Kim, 2019), while also highlighting the need for accessibility considerations across different participant groups.

Coaching topics were most self-selected and related to physical activity, nutrition, and sleep, consistent with previous findings (Young et al., 2020). Participants felt that choosing personally meaningful themes helped increase their motivation and could lead to secondary benefits in other areas. For example, improved sleep often led to greater physical activity, which in turn reinforced sleep quality. Coachees found health coaching to be a meaningful and motivating approach for making changes in their behaviors. The majority of coachees were either very satisfied or satisfied with the coaching they received, feeling that it supported them in achieving their goals and addressing the specific issues they sought help for. Similar results have been found in other studies examining coachees’ experiences with health coaching (Mitchell et al., 2021; Thom et al., 2015).

Coaches reported that coachees responded well to the coaching process. Noticeable behavioral changes often occurred within two to four sessions, particularly among coachees who were already somewhat ready for change. Motivation and internal reflection were seen as crucial to progress. Coaches played a key role in guiding clients toward small, manageable steps—an approach supported by Thom et al., 2015).

The importance of follow-up sessions was strongly emphasized by both coaches and coachees. The sense of accountability—knowing that one would report on progress— was described as motivating and encouraging (Kivelä et al., 2014). Some coachees used wearable devices to monitor their progress, which they found helpful and motivating, echoing the findings of Young et al. (2020).

The number of coachees per coach varied, and opinions on the ideal caseload differed. Balancing coaching responsibilities with regular healthcare duties was experienced as burdensome at times. Future programs should allow more time for preparation and delivery of coaching. Unlike in some previous studies (e.g., Nessen, 2014), role conflict between healthcare provider identity and coaching roles did not emerge in this study.

On the contrary, coaches felt their healthcare backgrounds enhanced their understanding of clients’ situations. This supports earlier research suggesting that nurses are well-suited to health coaching due to their communication and patient-centered care skills (Obro et al., 2023).

The training was effective in strengthening coaching competencies and confidence in using behavior change techniques, consistent with findings by Engelhard et al. (2018). Positive outcomes were achieved in this study within six months, but the long-term sustainability of these changes was not assessed. Dwinger et al. (2020) noted that some behavior changes reverted over time, suggesting that extended follow-up may be necessary to maintain changes.

In summary, coaches viewed the training positively but felt unable to fully absorb the amount of information presented during the short course or to develop sufficient readiness for practical coaching. Nevertheless, this uncertainty was not perceived by coachees, who experienced the coaching as supportive and motivating. They highlighted the coaches’ crucial role in creating a safe and encouraging atmosphere. Coachees felt they had gained practical tools for changing their behavior and new insights into how healthy habits could gradually become lasting lifestyle changes.

Through the coaching process, the coaches’ competence improved, and they emphasized that a background in healthcare provided an essential foundation for effective coaching, particularly in understanding clients’ health-related concerns and contexts. However, coaching was also perceived as an additional burden when performed alongside other healthcare responsibilities.

### Strengths and Limitations

A key strength enhancing the study’s credibility was its mixed methods design, which is particularly valuable in health sciences for providing deeper insights than single-method approaches. Survey data from participants were complemented and enriched by individual thematic interviews, allowing clarification and mutual understanding of responses. Additional strengths included a high survey response rate (79 %) and the comprehensive perspective achieved by capturing experiences from both coaches and participants.

However, some limitations must be acknowledged. One limitation of the study was the small sample size, as it consisted only of participants from a private medical center, which does not represent the broader population. These participants were predominantly middle-aged working women, many of whom lived alone or with a partner. As a result, this sample likely had fewer additional life challenges, such as unemployment or financial stress, which could affect their ability to participate in behavior change programs. This means the findings cannot be generalized to all population groups, particularly those who may be at higher risk for lifestyle-related diseases.

Another limitation was the dropout of two coaches during the pilot due to changing work schedules, which led to a reduction in the number of coaching sessions and coachees. This change in the coaching process may have influenced the overall results. Furthermore, although the study aimed to evaluate the impact of health coaching, the study’s duration (six months with seven sessions) may not have been long enough to assess the long-term sustainability of behavior changes. Future research should include follow-up assessments to examine whether the changes achieved in the study endure over time.

Despite the limitations, the findings of this study can be used in planning and implementation of future PrioMed^®^ health coaching training programs.

## Conclusions

The PrioMed^®^ health coaching training program is based on solid research evidence and shows promise in supporting sustainable behavior change. However, findings from the pilot highlight the need to strengthen practical coaching skills, particularly in real-life application and communication. Participants with healthcare backgrounds found the training more applicable, and coachees reported high satisfaction, viewing the coaching process as meaningful and supportive. These insights offer valuable guidance for developing future health coaching programs.

## Data Availability

All data produced in the present work are contained in the manuscript.

## REFERENCES

Carter A., Tamkin P., Wilson S. & Miller L. (2015). The Case for Health Coaching – Lessons Learned from Implementing a Training and Development Intervention for Clinicians across the East of England. Brighton: Institute of Employment Studies. https://www.employment-studies.co.uk/sites/default/files/resources/summarypdfs/heee0715a.pdf

Chew N.W.S., Ng C.H., Tan D.J.H., Kong G., Lin C., Chin Y.H., Lim W.H., Huang D.Q., Quek J., Fu C.E., Xiao J., Syn N., Foo R., Khoo C.M., Wang J-W., Dimitriadis G.K., Young D.Y., Siddiqui M.S., Lam C.S.P., Wang Y., Figtree G.A., Chan M.Y., Cummings D.E., Noureddin M., Wong V.W-S., Ma R.C.W., Mantzoros C.S., Sanyal A. & Muthiah M.D. 2023. The Global Burden of Metabolic Disease: Data from 2000 to 2019. A Cell Press Journal. 10.1016/j.cmet.2023.02.003

Clason C., Sterner-Stein K., Hirschman K.B., Barg F.K. & Riegel B. (2023). Developing effective health coaches: Experience gained in a clinical trial of a health coach intervention. Patient Education and Counseling 108 (2023) 107592 10.1016/j.pec.2022.107592

Davies F., Wood F., Bullock A., Wallace C. & Edwards A. (2020). Training in health coaching skills for health professionals who work with people with progressive neurological conditions: A realist evaluation. Health Expectations. 2020;23:919–933 10.1111/hex.13071

Dwinger S., Rezvani F., Kriston L., Herbarth L., Härter M & Dirmaier J. (2020). Effects of Telephone-based Health Coaching on Patient-reported Outcomes And Health Behaviour Change: A Randomized Controlled Trial. Plos One; 10.1371/journal.pone.0236861

Engelhard C., Lonneman W., Warner D. & Brown B. (2018). The implementation and evaluation of health professions students as health coaches within a diabetes self-management education program. Currents in Pharmacy Teaching and Learning 10 (2018) 1600–1608. 10.1016/j.cptl.2018.08.018

Finnish Institute for Health and Welfare, 2023. https://thl.fi/aiheet/kansantaudit/yleistietoa-kansantaudeista (Updated 5.12.2023)

Finnish National board on Research Integrity Tenk, 2023. The Finnish Code of Conduct for Research Integrity and Procedures for Handling Alleged Violations of Research Integrity in Finland. https://tenk.fi/sites/default/files/2023-05/RI_Guidelines_2023.pdf

Kappes M, Espinoza P, Jara V. & Hall A. (2023). Nurse-led telehealth intervention effectiveness on reducing hypertension: a systematic review. BMC Nursing. 10.1186/s12912-022-01170-z

Kim M. (2019). Effects of Customized Long-message Service and Phone-based Health-coaching on Elderly People with Hypertension. Iranian Journal of Public Health: Apr;48(4):655–663.

Kivelä K., Elo S., Kyngäs H. & Kääriäinen M. (2014). The effects of health coaching on adult patients with chronic diseases: A systematic review. Patient Education And Counseling. 10.1016/j.pec.2014.07.026

Langinkoski A., Hämäläinen J., Rapo K., Rinta M., Aroviita P. & Venojärvi M. (2024) Health Coaching 2.0: Redefining a Key Lifestyle Medicine Intervention, PrioMed Pilot Program Evaluation. MedRxiv. 10.1101/2024.06.10.24308710

Maini A., Fyfe M. & Kumar S. (2020) Medical students as health coaches: adding value for patients and students. BMC Medical Education (2020) 20:182. 10.1186/s12909-020-02096-3

McGill B., O’Hara B.J. & Phongsavan P. (2018). Participant Perspectives of a 6-month Telephone-Based Lifestyle Coaching Program. Public Health Research & Practise. 10.17061/phrp27451705

Miles L.M., Hawkes R.E. & French D.P. (2023). How the Behavior Change Content of a Nationally Implemented Digital Diabetes Prevention Program IS Understood and Used by Participants: Qualitative Study of Fidelity of Receipt and Enactment. Journal of Medical Internet Research. 10.2196/41214

Mitchell E.G., Maimone R., Cassells A., Tobin J.N., Davidson P., Smaldone A.M. & Mamykina L. (2021). Automated vs. Human Health Coaching: Exploring Participant and Practitioner Experiences. Proceedings of the ACM on Human-Computer Interaction. 10.1145/3449173

Napoles A.M., Santoyo-Olsson J., Chacon L., Steward A.L., Dixit N. & Ortiz C. (2019). Feasibility of a Mobile Phone App and Telephone Coaching Survivorship Care Planning Program Among Spanish-Speaking Breast Cancer Survivors. JMIR Publications; 10.2196/13543

Nessen T., Opava C.H., Martin C. & Demmelmaier I. (2014) From Clinical Expert to Guide: Experiences from Coaching People with Rheumatoid Arthritis to Increased Physical Activity. Physical Therapy Volume 94 Number 5 May 2014

Noll K., Wood A., Wood R. & Hebert E. (2022). Student Health Coaches Experiences with Adults with Chronic Health Conditions. American Journal of Health Studies: 36(4) 2022

Nordic Health Academy. (2024) PrioMed-research project: https://nha.fi/priomed-tutkitusti-toimivaa-elintapavalmennusta-tyoyhteisoille/ (20.03.2024)

Obro L.F., Osther P.J.S., Ammentorp J., Pihl G.T., Korgh P.G. & Handberg C. (2023). Healthcare Professionals’ Experiences and Perspectives of Facilitating Self-Management Support for Patients with Low-Risk Localized Prostate Cancer via mHealth and Health Coaching, International Journal of Environmental Research and Public Health 2023, 20, 346. 10.3390/ijerph20010346

Olsen J.M. (2014). Health Coaching: A Concept Analysis. Nursing Forum.10.1111/nuf.12042

Piercy K.L., Troiano R.P., Ballard R.M., Carlson S.A., Fulton J.E., Galuska D.A., George S.M. & Olson R.D. 2018. The Physical Activity Guidelines for Americans. Journal of the American Medical Association. 10.1001/jama.2018.14854

Pounds K., Offurum A. & Moultry A.M. (2015). First year pharmacy students as health coach in the management of hypertension. Pharmacy Education, 2015; 15 (1) 111 – 115

Thom D.H,. Hessler D., Willard-Grace R, DeVore D., Prado C., Bodenheimer T. & Chen E.H. (2015). Health Coaching by Medical Assistants Improves Patients’Chronic Care Experience. The American Journal of Managed Care Oct;21(10):685–91. https://www.ajmc.com/view/health-coaching-by-medical-assistants-improves-patients-chronic-care-experience

Wharton S., Lau D.C.W., Vallis M., Sharma A.M., Biertho L., Campell-Scherer D., Adamo K., Alberga A., Bell R., Boulé N., Boyling E., Brown J., Calam B., Clarke C., Crowshoe L., Divalentino D., Forhan M., Freedhoff Y., Gagner M., Glazer S., Grand C., Green M., Hahn M., Hawa R., Henderson R., Hong D., Hung P., Janssen I., Jacklin K., Johnson-Stoklossa C., Kemp A., Kirk S., Kuk J., Langlois M-F., Lear S., McInnes A., Macklin D., Naji L., Manjoo P., Morin M-P., Nerenberg K., Patton I., Pedersen S., Pereira L., Piccinini-Vallis H., Poddar M., Poirier P., Prud’homme D., Salas X.R., Rueda-Clausen C., Russel-Mayhew S., Shiau J., Sherifali D., Sievenpiper J., Sockalingam S., Taylor V., Toth E., Twells L., Tytus R., Walji S., Walker L. & Wicklum S. (2020). Obesity in Adults: A Clinical Practise Guideline. Canadian Medical Association Journal. 10.1503/cmaj.191707

Wolever R.Q., Simmons L.A., Sforzo G.A., Dill D., Kaye M., Bechard E.M., Southard M.E., Kennedy M., Vosloo J. & Yang N. (2013). A Systematic Review of the Literature on Health and Wellness Coaching: Defining a Key Behavioral Intervention in Healthcare. Global Advances in Health and Medicine. 10.7453/gahmj.2013.042

World Health Organization. (2021). Implementing telemedicine services during COVID-19: guiding principles and considerations for a stepwise approach. https://iris.who.int/bitstream/handle/10665/336862/WPR-DSE-2020-032-eng.pdf?sequence=5 (Updated11.3.2024)

World Health Organization. (2022). Noncommunicable diseases. https://www.who.int/news-room/fact-sheets/detail/noncommunicable-diseases (Updated 16.09.2023)

World Health Organization (2023). Advancing the global agenda on prevention and control of noncommunicable diseases 2000 to 2020. Looking forward to 2030. https://iris.who.int/bitstream/handle/10665/370425/9789240072695-eng.pdf?sequence=1 (05.03.2024)

Young H.M., Miyamoto S., Dharmar M. & Tang-Feldman Y. (2020). Nurse Coaching And Mobile Health Compared with Usual Care to Improve Diabetes Self-efficacy for Persons with Type 2 Diabetes: Randomized Controlled Trial. JMIR Mhealth Uhealth; 10.2196/16665

